# Noninvasive Proteomic Markers for Respiratory Tract Infections in Mechanically Ventilated Patients

**DOI:** 10.1101/2022.08.17.22278888

**Authors:** Dapeng Chen, Marek A. Mirski, Shuo Chen, Alese P. Devin, Caroline R. Haddaway, Emily R. Caton, Wayne A. Bryden, Michael McLoughlin

## Abstract

**Introduction:** Early and accurate diagnosis of respiratory tract infections (RTI) in critical care settings is essential for appropriate antibiotic treatment and lowering mortality. The current diagnostic methods face critical challenges, including the lack of noninvasive specimens from the site of infection and molecular biomarkers that can predict disease progression and treatment effect. In this study, we addressed these critical challenges by developing a noninvasive method based on the characterization of truncated proteoforms contained in exhaled air collected from mechanically ventilated patients.

**Methods:** Exhaled air samples were collected from twenty-five intubated patients with RTI and twenty-two intubated patients without RTI, determined by clinical data and microbiological testing. Truncated proteoforms were identified using top-down proteomics. Feature selection algorithms were used to identify significant truncated proteoforms associated with RTI. A score system combining the significant truncated proteoforms was constructed and evaluated using multiple logistic regression to predict RTI.

**Results:** The results showed that six truncated proteoforms of lung structure and proteolytic proteins were statistically different between intubated patients with and without RTI. Specifically, the truncated proteoforms of collagen type VI alpha three chain protein, matrix metalloproteinase 9, and putative homeodomain transcription factor 2 were found to be independently associated with RTI. A score system named TrunScore was constructed by combining the three truncated proteoforms, and the diagnostic accuracy was significantly improved compared to individual truncated proteoforms.

**Conclusions:** In this study, we presented a noninvasive method to address the current challenges in diagnosing RTI in critical care settings, by characterizing truncated proteoforms contained in exhaled air from intubated patients. The method provides an accurate prediction for RTI in mechanically ventilated patients and can help diagnose other respiratory tract diseases.

## INTRODUCTION

Mechanically ventilated patients are at a higher risk of acquiring respiratory tract infections (RTI) in intensive care units (ICU) [1]. A worldwide study investigating 7,087 infection cases in ICU demonstrated that 4,503 cases (64%) were respiratory in nature [1]. RTI in intubated patients leads to pneumonia, sepsis, and septic shock, resulting in high mortality and increased hospital cost [2]. The current management strategy for intubated patients with RTI is to initiate broad-spectrum antibiotics as soon as patients show clinical signs of RTI [2]. Since specific conventional microorganisms cause most hospital associated RTI, administration of broad-spectrum antibiotics typically yields good treatment outcomes, and therefore is encouraged [2]. As a result, intubated patients contribute to major antibiotic prescriptions in critical care settings, a primary source driving antibiotic resistance [3].

The lack of a noninvasive method to secure reliable pulmonary-based inflammatory material, or molecular biomarkers, is a primary barrier to rapidly diagnosing RTI in critical care settings, especially in patients on mechanical ventilators [2]. The current diagnostic strategy relies on non-specific clinical observations, such as the presence of tracheal secretions, chest X-ray findings consistent with pneumonia, elevated body temperature or white blood cell counts, or diminished oxygenation capacity [2]. Scoring systems, such as clinical pulmonary infection score (CPIS), have been developed based on these clinical symptoms [4]. Although the clinical findings and scoring systems can be used to suggest antibiotic treatment, they generally lack sensitivity and specificity for RTI diagnosis, thereby making it challenging for clinicians to provide rational clinical decisions regarding treatment [5]. Quantitative microbial culture of specimens collected from the lower respiratory tract, such as the noninvasive endotracheal aspirate (ETA), have been used for RTI diagnosis, but face a critical challenge in providing essential information on whether the identified bacteria result from common respiratory tract colonization or are the etiology of the infection [2,6]. Bronchoalveolar lavage (BAL) has been used as a high-quality specimen collection technique from the lower respiratory tract for causative diagnosis in intubated patients. However, this method is invasive, and is most commonly performed only when other evaluations are inconclusive, and there is a determined need for a specific diagnosis [2]. Due to these limitations, over 50% of patients administrated in intensive care units were treated without an appropriate diagnosis [7]. Thus, the difficulty of obtaining samples from the site of infection, and the absence of accurate diagnostic molecular biomarkers, limit the current diagnosis, pathogen identification, and management of RTI in intubated patients. There is an urgent need to develop a noninvasive method for sampling the source of infection and discovering accurate molecular biomarkers for RTI diagnosis.

Mounting evidence demonstrates that human exhaled air contains non-volatile organic compounds (NOCs) of medical diagnostic potential [8-11]. Previously we developed a device, BreathBiomics™, for the noninvasive collection of proteins and peptides in human exhaled air based on surface chemistry [12]. The design of BreathBiomics™ addressed several critical challenges in capturing NOCs contained in human exhaled air, such as low capture efficiency and lack of feasibility for clinical use [12]. Our initial proteomic analysis demonstrates that the proteins identified in the exhaled air overlapped with BAL and suggests it can be used as a noninvasive collection method for lung proteins [12]. Proteases are a class of enzymes that catalyze the cellular proteolysis process. As a critical component of the self-defense system, proteases are upregulated during infections and initiate protein hydrolysis by breaking down intact proteins into small peptides, called protein fragments or truncated proteoforms. The cleavage of substrates, including lung structure proteins, cytokines, and immunoglobins, has been observed in lung diseases associated with chronic inflammation and infections [13,14]. Unlike other dynamic protein translational modifications such as phosphorylation and glycosylation, protein hydrolysis is irreversible, promoting them as ideal biomarker candidates to indicate pathological lung changes. In this study, we hypothesized that the truncated proteoforms, an end-point product of activated proteases in bacterial infection, can be captured from the exhaled air from intubated patients using the noninvasive BreathBiomics™ and offered prediction values for RTI.

## MATERIALS AND METHODS

### Mechanically Ventilated Patient Information

All study participants had been provided with written informed consent. The study was approved by the institutional review boards at The Johns Hopkins University School of Medicine (Application number: IRB002494495). All tests presented in this work were conducted in accordance with the approved protocols. In this work, 30 intubated patients in the neurological ICUs at The Johns Hopkins Hospital, and 47 exhaled air samples were collected (Table 1). Age, gender, race, ethnicity, primary diagnosis, medication, sample collection time, microorganism identification information, white blood cell test, body temperature, fraction of inspired oxygen (FiO2) test, and pulmonary radiography were provided (Table 1). The microorganism identification was conducted in the clinical laboratory at The Johns Hopkins Hospital and interpreted by the ICU clinicians. The positive respiratory tract infection was diagnosed by the physicians at The Johns Hopkins Hospital using clinical criteria and confirmed when the tract samples, including sputum, endotracheal tube sample (ET), or bronchoalveolar lavage (BAL), were cultured positive in the clinical laboratory at The Johns Hopkins Hospital.

**Table 1.** Patient characteristics in this study.

| Characteristic | All Patients<br>n = 30 | Patients without RTI<br>n = 17 | Patients RTI<br>n = 13 |
| --- | --- | --- | --- |
| Age (Years) | 61.2 (19-84) | 64.5 (29-84) | 57.3 (19-74) |
| Gender (F, M) | (F)13, (M)17<br>(F)43.3%, (M)56.7% | (F)9, (M)8<br>(F)47.1%, (M)52.9% | (F)4, (M)9<br>(F)30.8%, (M)69.2% |
| White Blood Cell Counts,<br>x 10<sup>3</sup> per cubic millimeter | 12.4 (6.9-26.6) | 11.7 (6.9-26.6) | 13.0 (7.7-21.0) |
| Body temperature (°C) | 37.1 (35.3-38.5) | 37.2 (36.0-38.5) | 37.0 (35.3-37.9) |
| fiO<sub>2</sub> % | 41.6 (30-100) | 40 (30-50) | 42.9 (30-100) |

### Collection of Exhaled Air in Intubated Patients

For the exhaled air collection, BreathBiomics™, developed in our previous study, was connected to a tee fitting which was installed in the exhaust tubing on the mechanical ventilator (Figure 1A-C) [12]. The collection column is connected to a CO2 sensor (Gas Sensing Solutions Ltd, Cumbernauld, UK) and a mini diaphragm pump (Parker Hannifin Corporation, Cleveland, OH). The flow rate of the pump is set up to 0.5 liter/minute. The CO2 sensor is used to record individual exhaled CO2 level in the exhaust tubing on the mechanical ventilator. Once the collection was completed, BreathBiomics™ was removed from the ventilator and decontaminated in the clinical laboratory at The Johns Hopkins Hospital before sent to the laboratory at Zeteo Tech for further sample preparation (Figure 1C). To elute non-volatile organic molecules, 300 µL of 70% acetonitrile (v/v, in water, Thermo Fisher Scientific, Waltham, MA) was added in the column and pushed through using a syringe. The organic solvent was removed by overnight lyophilization. Then, 20-50 µL of 0.05% trifluoroacetic acid (v/v, in water, Thermo Fisher Scientific) was added. The samples were saved in the laboratory refrigerator for further analysis.

**Figure 1.**
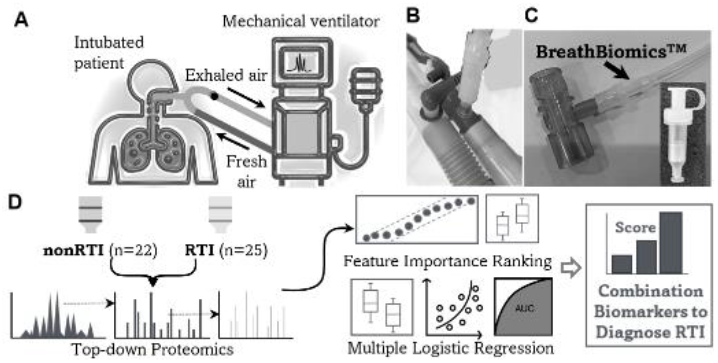
The overall workflow for the collection of truncated proteoforms from the exhaled air in intubated patients and data analysis. (A) The configuration of the exhaled air collection in patients on mechanical ventilators. The red dot indicates where the air collection column is installed. (B) The position of the tee fitting, indicated by the dark arrow, on the exhaled air tubing of the mechanical ventilator. (C) the position of the collection column, indicated by the dark arrow, on the tee fitting and the collection column after sample collection. (D) The workflow of data analysis to identify significant truncated proteoforms to predict patients with RTI.

### Identification of Truncated Proteoforms Using Top-down Proteomics

Identification of truncated proteoforms was conducted using the workflow established in our previous study [15]. Generally, 18 µL of exhaled air sample was injected into a an EASY-nLC 1000 system (Thermo Fisher Scientific, Waltham, MA). For high-performance liquid chromatography, the sample was loaded in an Acclaim PepMap 100 C18 trap column (0.2 mm×20 mm, Thermo Fisher Scientific) at the flow rate of 5 µL/min and separated using an Acclaim PepMap 100 analytical column (75 µm×150 mm, Thermo Fisher Scientific) at the flow rate of 300 nL/min from 5-70% of 80% acetonitrile and 0.1% formic acid (v/v, in water) for 60 minutes. Precursor ions and fragmentation ions generated from collision-induced dissociation were acquired in a LTQ orbitrap mass spectrometer (Thermo Fisher Scientific) at the mass resolution of 60,000. The raw files generated from the mass spectrometry were loaded in MaxQuant software (maxquant.org/) for the identification of truncated proteoforms in exhaled air samples. We used defaulted parameters for the database searching expect that the enzyme option was set to “nonspecific” and no fixed modification was defined. Human proteome database was constructed using UniProtKB/Swiss-Prot database (uniprot.org) which included 20,387 reviewed *Homo sapiens* protein entries as of the data of manuscript submission.

### Data Analysis and Statistics

Previously we established a workflow to identify features of statistical significance for the sample type classification, including mass spectrometric data processing, feature selection, and logistic regression to predict a binary outcome using multiple mass spectrometric features [10,11]. Generally, mass spectrometric features were normalized by total ion chromatography. Data transformation, scaling, and centering was conducted by using the log transformation method previously described in our studies [10,11]. To select the most significant features that contribute to the classification between patients without RTI and with RTI, a ranking algorithm for omics, Significance Analysis of Microarrays (SAM), was used [10,11]. SAM provides featuring ranking based on each feature’s statistics and fold-change. A two-tailed t-test was also applied to all the truncated proteoforms identified in this study to acquire raw p values, which were then adjusted by the application of false-discovery rate (FDR) using the Benjamini-Hochberg method.

Multiple logistic regression was used to evaluate the association between the RTI status and measured variables, including the patients’ characteristics and truncated proteoforms with statistical significance. Receiver operating characteristic curves (ROC) and area under the curve (AUC) were constructed using significant features after p-value adjustment using the logistic regression model (Figure 1D). We created a score model based on the truncated proteoforms with statistical significance to evaluate the performance of the combined truncated proteoforms for separating patients without RTI and with RTI. In this model, the cutoff values for truncated proteoforms were generated based on the specificity and sensitivity values constructed in ROC curves, and a point is provided per truncated proteoform when a value is larger than the cutoff value. This score system was evaluated using multiple logistic regression, and the performance was provided using AUC (Figure 1).

## RESULTS

### Truncated Proteoform Profiles Identified in The Exhaled Air Collected from Intubated Patients

In total, 263 truncated proteoforms of 80 proteins were identified in the exhaled air samples collected from the intubated patient (Figure 2A). The distribution of truncated proteoforms in each sample showed more truncated proteoforms identified in samples of intubated patients with RTI (Figure 2B). Statistical analysis showed this difference in identified truncated proteoforms was significant (Figure 2C). In samples collected from intubated patients with RTI, approximately 125 truncated proteoforms were identified, in contrast to approximately 55 in intubated patients without RTI (Figure 2C).

**Figure 2.**
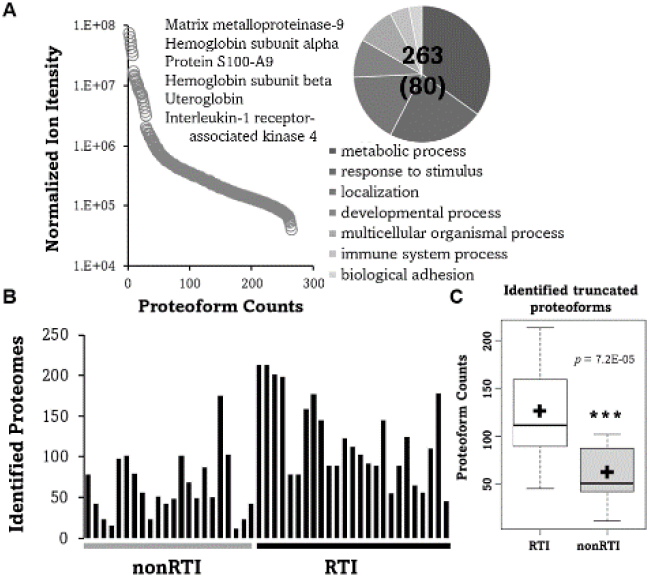
Truncated proteoform profiles in exhaled air collected from intubated patients. (A) Distribution of mass spectrometric features of truncated proteoforms and protein function analysis. (B) Counts of truncated proteoforms in intubated patients without or with infection (RTI). (C) Statistical analysis of truncated proteoform counts between the two groups.

Representative ion fragmentation maps of three truncated proteoforms of S100-A9, interleukin-1 receptor-associated kinase 4, and uteroglobin were demonstrated for the confident identification of truncated proteoforms, including the identification of two sites of methionine oxidation on the proteoform of uteroglobin (Figure 3A-C).

**Figure 3.**
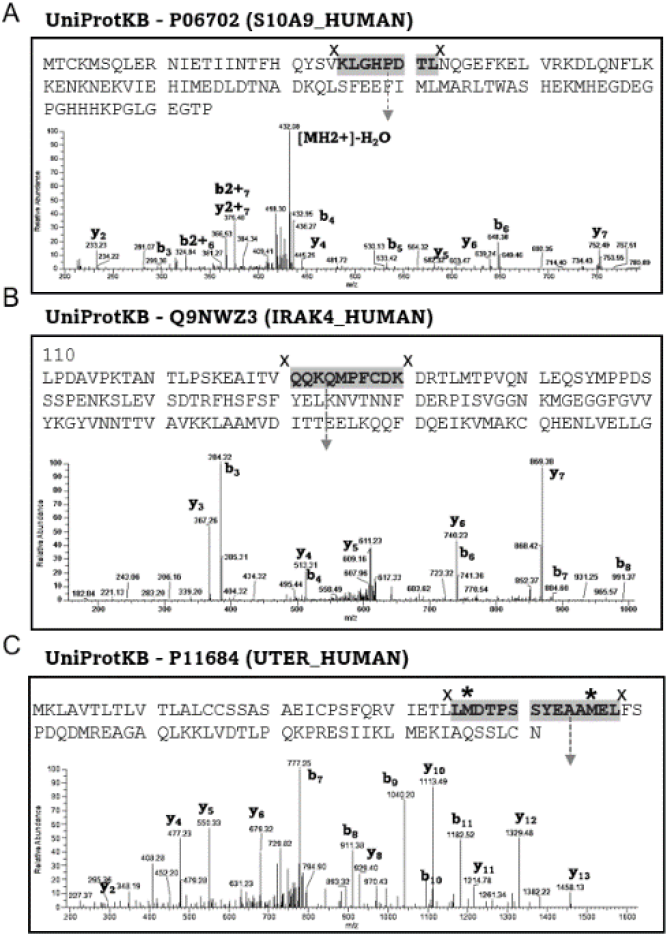
Representative ion fragmentations of truncated proteoforms and demonstration of residues carrying the cleavage sites of truncated proteoforms. (A-C) Ion fragmentations of 3 representative truncated proteoforms.

### Identification of Truncated Proteoforms Contributing to The Separation Between NonRTI and RTI Samples

SAM analysis and the Benjamini-Hochberg method were used to identify truncated proteoforms contributing to the separation between nonRTI and RTI groups (Table 2, Figure 4). Both methods provide statistical significance analysis with the FDR adjustment and can be used for feature reduction. Besides, SAM analysis provides feature importance ranking based on the features’ separation power. Six truncated proteoforms, CO6A3 (2781-2792), MMP9 (673-691), PHTF2 (271-285), IRAK (121-130), CYTA (2-17), and DEN2B (628-637), were found to be significantly different between two groups. Five truncated proteoforms, except PHTF2 truncated proteoform, were found to be higher than in the RTI group (Table 2, Figure 4). SAM analysis ranked all the 6 truncated proteoforms, and CO6A3 was found to be the top feature in the list (Figure 4). After FDR adjustment at 0.05, all 6 truncated proteoforms showed statistical significance between the two groups (Table 2).

**Table 2.**
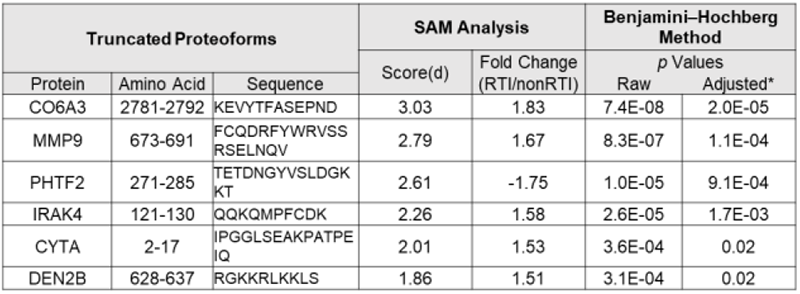
The fold-change and statistical values of 6 truncated proteoforms revealed by SAM analysis and Benjamini-Hochberg method.

**Figure 4.**
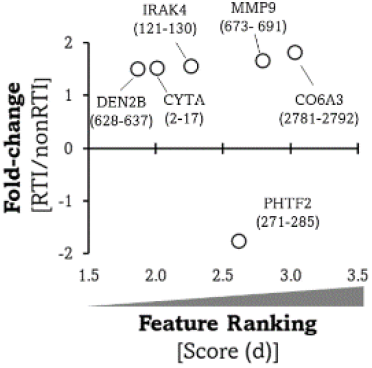
SAM analysis results. (A) Distribution of feature ranking scores and fold-changes of 6 significant truncated proteoforms in SAM analysis.

### Accuracy of 6 Truncated Proteoforms in Predicting RTI

The accuracy of the 6 truncated proteoforms was evaluated by calculating AUC values in ROC curves. The results showed that CO6A3 truncated proteoform provided the highest discriminative value with AUC of 88.5%, followed by MMP9 (79.3%), PHTF2 (76.5%), CYTA (69.1%), DEN2B (62.2%), and IRAK4 (62.0%) (Figure 5). To evaluate the prediction of RTI using the 6 truncated proteoforms, multiple logistic regression was conducted to variables, including patients’ clinical outcomes and identified proteoforms. The results showed that 3 truncated proteoforms of CO6A3, MMP9 and PHTF2 were significantly associated with RTI (Table 3). The most significant truncate proteoform is MMP9 with a *p* value of 0.006 (Table 3).

**Table 3.**
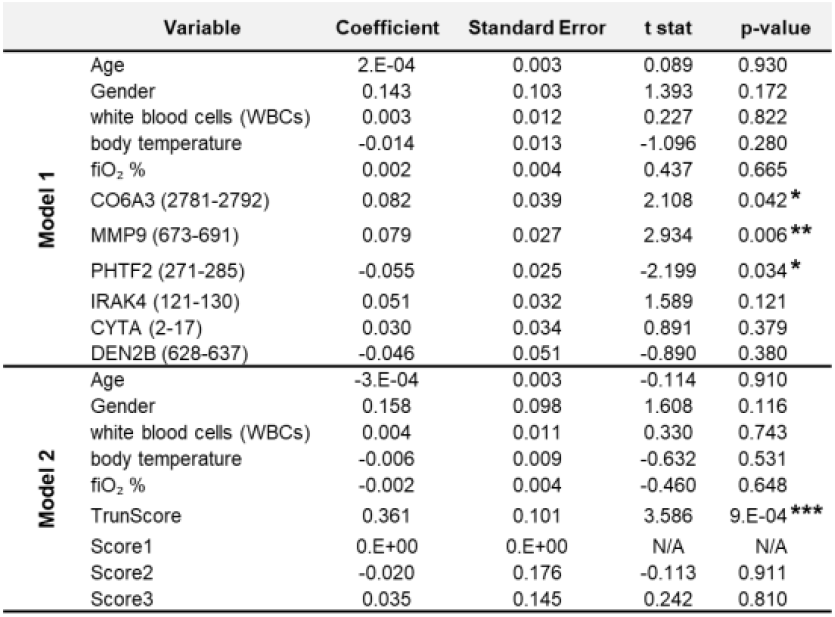
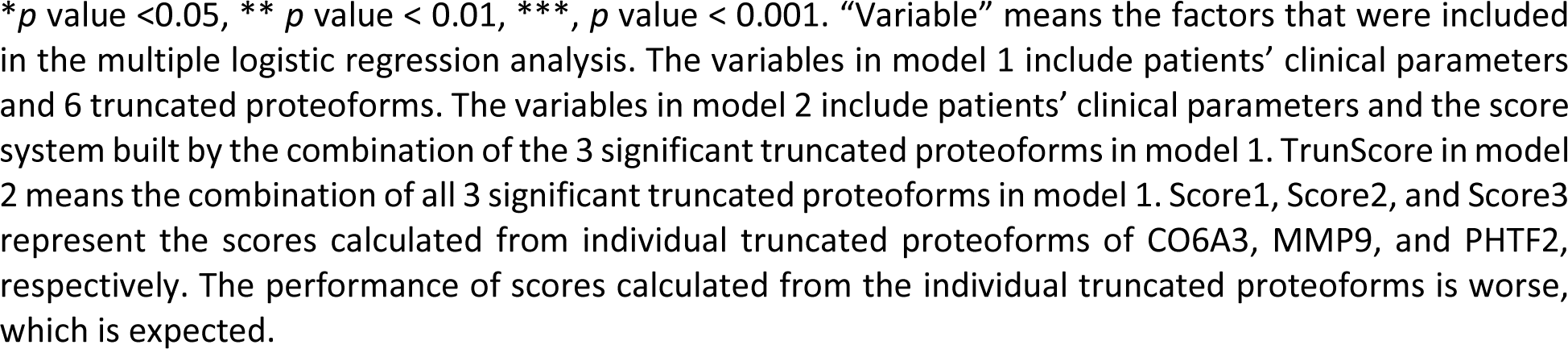
Multiple logistic regression analysis of variables used for separating between patients without RTI and with RTI in the two models.

**Figure 5.**
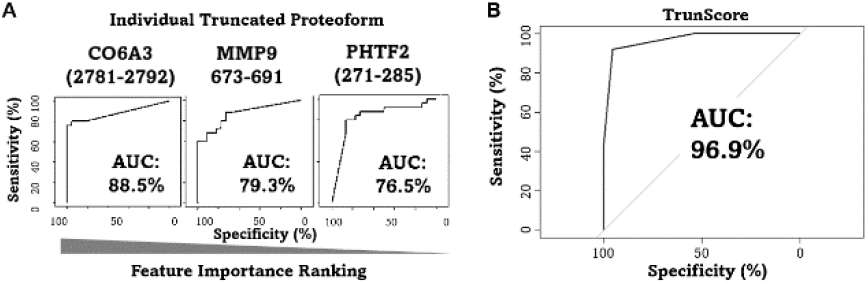
The performance of different models in the separation between two groups. (A) ROC curves and AUC values for individual truncated proteoforms in separating between patients without RTI and with RTI. (B) The ROC curve and the AUC value of the score system in separating patients without RTI and with RTI.

### Combination of Truncated Proteoforms into A Score System to predict RTI

A combination of several biomarkers is commonly used to improve a model’s diagnostic performance. In this study, we evaluated whether combining truncated proteoforms would result in a better discriminative value in the prediction of RTI. For this purpose, the 3 truncated proteoforms that were found to be significantly associated with RTI in the multiple logistic regression analysis, truncated proteoforms of CO6A3, MMP9, and PHTF2, were given a score of 0 or 1 depending on the cutoff threshold determined in the ROC curves. For example, a sample scores 0 if the mass spectrometric value of a truncated proteoform measured in this sample is below the cutoff value and a score 1 if above the cutoff values. Based on this concept, the TrunScore of each sample ranges between 0 and 3. 0 means all three truncated proteoforms are under the cutoff value, and 3 means all three truncated proteoforms are above the cutoff value.

The multiple logistic regression analysis and the performance of TrunScore using AUC in ROCs showed that the TrunScore provided a much better discriminative value than individual truncated proteoforms in predicting RTI (Table 3, Figure 5). ROC curves showed the AUC was 96.9%, compared to 88.5% provided by the best marker CO6A3 truncated proteoform (Figure 5A, B). The *p*-value calculated in multiple logistic regression analysis when using individual truncated proteoforms is 0.006 for MMP9 (Model 1, Table 3). The *p*-value is small than 0.001 when using the TrunScore in the model 2 (Table 3). To determine the best score for RTI diagnosis, confusion matrices using different cutoff scores were constructed. The cutoff score of 2 provided the best accuracy (93.6%), precision (95.8%), and recall (92.0%) values.

## DISCUSSION

The most critical finding in this hypothesis-driven study is that in using a noninvasive method, truncated proteoforms contained in exhaled air can be collected from intubated patients and used to predict RTI. We showed that the truncated proteoform profiles differed between intubated patients with and without RTI. We identified six truncated proteoforms significantly associated with RTI and created a better prediction model using a score system that combined the significantly truncated proteoforms. Our findings provide a novel methodology to address the current challenges in the diagnosis of RTI in intubated patients.

The urgent need for a noninvasive method for diagnosing bacterial infections in critical care sites has drawn remarkable attention in recent years [16-19]. The current specimen collection methods are invasive and pose a major barrier to a better characterization of RTI [2]. The development of a noninvasive sampling method has several potential benefits. The noninvasive method enables repeated sampling without adding risk in critically ill patients, so that a disease trajectory can be constructed. The direct sampling from the lower respiratory tract would offer specimens that better represent the site of infection and thus provide a better specificity for the diagnosis, which is a limitation of using biomarkers derived from peripheral blood samples that cannot offer localization to specific targets of inflammation and infection. The noninvasive sampling method, easy and risk-free, would further encourage patients to enroll in clinical trials that can be beneficial to therapeutic and diagnostic research.

Human breath and exhaled air have the promise to be used as a noninvasive source in clinical use [8-11]. Organic molecules contained in human breath and exhaled air have been studied to develop a noninvasive method for detecting lung disease exacerbation and infections [8, 20]. The organic molecules in human breath include two main types: volatile organic compounds (VOCs) and NOCs [21]. VOCs are gas molecules that can be emitted from non-biological sources, such as diets, plants, and home cleaning products, and thus lack specificity for biomarker use [22]. On the contrary, NOCs are large molecules that exclusively originate from organisms, either humans or pathogens and thus are more suitable to be used as surrogate biomarkers.

Noninvasive sampling methods targeting NOCs have been developed for use in clinical settings. McNeil et al. reported using inline heat moisture exchange (HME) filters to collect proteins from patients with acute respiratory distress syndrome (ARDS) [8]. HMS filter is a standard piece installed on mechanical ventilators where exhaust air is present. The authors hypothesized that proteins could be captured on the HMS filter as exhaled breath condensate emitted from lower airways. For this purpose, undiluted pulmonary edema fluid (EF) samples were collected, and the protein profiles acquired from EF samples were used to compare with HEM fluid samples [8]. The results showed a similar protein profile between the two types of samples and suggested that HME could be a noninvasive alternative to EF for distal sampling airspace in patients with ARDS [8].

The protein profiles between McNeil’s and our study overlapped, and some major findings are consistent between the two studies. The protein profiling showed that proteins of lung structure and inflammatory responses were observed in both studies, including filamin A, actin, MMP9, myosin-9, fibronectin, and the S100 protein family. Those proteins were also reported in BAL samples collected from intubated patients, suggesting that both HME filters and BreathBiomics™ can capture proteins of lower airways, and proteins associated with inflammatory response were of great potential to be biomarkers. In addition, McNeil reported that the total protein was significantly higher in ARDS patients [8]. In our study, the identified proteoforms were significantly higher in intubated patients with RTI. This consistency is very interesting, considering the two studies using different proteomic profiling approaches in different cohorts. The activation of inflammatory response could explain the higher protein content in intubated patients with ARDS, as McNeil showed higher levels of MMP9 and myeloperoxidase. In our study, the elevated truncated proteoforms in intubated patients with RTI could directly result from protease/antiprotease imbalance. Nevertheless, the two studies strengthened the concept of using breath air and exhaled air for noninvasive methods to address the current challenges in detecting lung diseases and infection.

Several advantages of our approach should be noted. Although McNeil’s study demonstrated the use of HEM filters to replace invasive EF fluid collection, the capture mechanism is neither investigated further nor optimized [8]. Several types of HEM filters are used on mechanical ventilators. The essential components are the same, as they are made of sponge-like materials with hygroscopic properties. It is speculated that the capture of proteins is mainly determined by condensation. The primary issue of this “condensation capture mechanism” is that submicron particles cannot be efficiently collected. Indeed, a recent study demonstrated that HEM filters were not useful in detecting SARS-CoV-2 viruses in intubated patients [23]. On the contrary, the capture mechanism of BreathBiomics™ relies on the molecular interactions between the immobilized chemical functional groups and proteins [12]. Our previous studies showed high capture efficiency in proteins contained in submicron particles [12]. Since the particles in human breath and exhaled air are mainly within the submicron size range, BreathBiomics™ is positioned to be more effective in collecting infection-related proteins in intubated patients. Another critical challenge in human breath analysis is sample normalization. We addressed this limitation by enabling the recording of individual CO2 levels for data normalization, which has not been reported in previous studies using human breath.

In this study, we conducted a top-down proteomics strategy to characterize truncated proteoforms in the exhaled air samples collected from the intubated patients. Bottom-up and top-down proteomics-based protein profiling is routinely used for biomarker discovery research [24]. The major difference between the two approaches is protein data interpretation. Since proteins are digested in the bottom-up proteomics, protein information and quantitative analysis are not straightforward as the information need to be back-tracked, which could cause loss of information [24]. Top-down proteomics overcomes this limitation as protein quantitation can be directly applied to the proteoforms present in biological samples [25, 26]. Truncated proteoforms can be characterized with bottom-up proteomics to identify the N- and C-terminal residues, which provides comprehensive information about the proteases involved [27]. However, this method cannot reveal the entire length and identification of the truncated proteoforms.

In this study, we demonstrated that the truncated proteoforms, characterized by top-down proteomics, can be used to differentiate intubated patients with RTI. Unlike traditional methods, which rely on bacterial culture, the identification strategy is based on host-response molecular markers. Our first observation is that more truncated proteoforms were identified in intubated patients with RTI, presumably due to excessive protease activation during bacterial infections. Indeed, neutrophil proteases were reported to be significantly elevated in patients with ventilator-associated pneumonia [14]. Wilkinson, Li, and Hellyer reported that neutrophil elastase, MMP8, and MMP9 were significantly higher in BAL samples collected from patients with VAP than in non-VAP cases [13,14,28]. The amino acid residues of the truncated proteoforms’ N- or C-terminus were respectively characterized in nonRTI and RTI groups. In both groups, the major residues identified are serine, valine, and lysine for the N-termini, and arginine, lysine, leucine, and serine for the C-terminus. Since serine proteases, such as neutrophil elastase (NE), preferentially cleave glycine, serine, valine, leucine, and alanine at the C-terminus, it can be interpreted that the truncated proteoforms constitute a significant result of NE protease activation. Previous studies suggest that multiple proteases contribute to the production of truncated proteoforms [26,27]. Therefore, further research combining the advantages of bottom-up and top-down proteomics should be conducted to characterize the proteases activated during respiratory tract infections fully.

We showed that the mass spectrometric intensities of 6 truncated proteoforms significantly differed between the nonRTI and RTI groups. Among the 6 truncated proteoforms, 2 proteins, CO6A3 and CYTA, are lung structure proteins, and 3 proteins, MMP9, IRAK4, and DEN2B, have protease/anti-protease function. The evaluated truncated proteoforms of lung structure and proteolytic enzymes indicate that the balance between proteases and anti-proteases plays a vital role in the pathogenesis of respiratory tract infections. To the best of our knowledge, we demonstrated for the first time that the characterization of truncated proteoforms could reveal the pathological processes during respiratory tract infections. In our previous study, we established a feature selection algorithm, SAM, to evaluate the p values from the statistical analysis [10,11]. The features that show higher ranking scores calculated in SAM analysis, theoretically the ones with the lowest *p* values and highest fold-changes, are more easily identified in top-down proteomics and potentially biologically more relevant. We employed a multiple logistic regression model to evaluate the truncated proteoforms for predicting respiratory tract infections. When combining the two analysis results, 3 truncated proteoforms of CO6A3, MMP9, and PHTF2 turned out to be the most discriminative features for the prediction of RTI. Since combining biomarkers to improve diagnostic performance is a routine practice to predict the outcome, we created a model in which the 3 significant truncated proteoforms were combined into a TrunScore. Although the TrunScore is simply calculated from the cutoff thresholds in ROC curves, it provides very clear information which can likely assist the clinicians’ decision besides other clinical notes.

The methods for distinguishing viral from bacterial respiratory tract infections are urgently needed [29]. No patients with viral respiratory tract infections were included in this study. Thus, future studies should focus on investigating if the characterization of truncated proteoforms can be applied to differentiate between bacterial and viral cases, which has been a critical challenge for clinicians in critical care settings. Studies of characterizing proteases and anti-proteases in viral infections are not as common as in bacterial cases, except for COVID-19 [30]. Nevertheless, it has been reported that human trypsin-like serine proteases are primarily associated with the cleavage of hemagglutinin proteins in influenza infections [31]. The protease-antiprotease imbalance was investigated in serum and BAL samples collected from patients with severe COVID-19, and the results showed that MMP12 and NE were elevated in serum but not in BAL samples in COVID-19 patients [30]. Another study using nasal swab samples reported the dominant presence of truncated proteoforms in COVID-19 patients [32]. Castillo et al. performed immunoassays to characterize pulmonary surfactant proteins in plasma and airways samples [32]. The results showed that degradation of surfactant protein D could be used to predict the progression of acute respiratory distress syndrome in ventilated patients with COVID-19. Based on this evidence, the truncated proteoform profile can be used to differentiate viral from bacterial infection cases.

Several limitations in our study need to be noted. Although protein profiling analysis demonstrated that proteins of truncated proteoforms showed good overlap with reported BAL proteomes, a parallel comparison in which BAL and exhaled air samples are collected from the same intubated patient needs to be conducted. The best scenario would be the comparison in protein profiles conducted in BAL, HEM fluid, and exhaled air. In addition, the current analysis was based on mass spectrometric intensities, and absolute quantitative measurement of truncated proteoform concentrations in exhaled air needs to be conducted for clinical use. For example, although the identified truncated proteoforms were significantly higher in patients with RTI, a cutoff value cannot be calculated based on mass spectrometric analysis because of differences in mass spectrometry instruments. Lastly, this study constructed a Scoring system based on a collection of breath samples in a single ICU setting, and no forward validation has yet to confirm the results herein.

## Conclusion

In conclusion, we addressed the current critical challenges in diagnosing respiratory tract infections in critical care settings by developing a noninvasive method and a score system based on the profiles of truncated proteoforms contained in exhaled air. The individual truncated proteoforms show significant differences in intubated patients with respiratory tract infections, and the diagnostic accuracy improved when a score system combined the truncated proteoforms. Continuous studies should focus on validating truncated proteoforms in different critical care settings and in prospective trials and develop the workflow so the current strategy can assist clinical decision-making for the early management of patients with suspected respiratory tract infections.

## Data Availability

All data produced in the present study are available upon reasonable request to the corresponding author.

## Availability of data and materials

Please contact the corresponding author for data use.

## Authors contributions

DC, WAB, and MM designed the study. DC designed the experiments. DC and SC analyzed the date. APD, CRH, and ERC collected and processed samples that were received from The Johns Hopkins Hospital. MAM was responsible for exhaled air samples and maintained patient database. All authors agreed with the results. All authors approved the manuscript.

